# Undergraduate students with and without mental health concerns have different perceptions of disclosing mental health challenges to instructors

**DOI:** 10.1101/2024.11.25.24317913

**Authors:** J. Michael Sizemore, Bailey Von der Mehden, Elisabeth E. Schussler

## Abstract

A significant proportion of undergraduates report having mental health concerns (MHC), which are associated with reduced academic success. Students with MHC are encouraged to seek help from their instructors but may not because of perceived negative reactions by instructors and peers. This suggests stigma about MHC may differentially impact disclosure perceptions of students with MHC compared with their peers, yet the perceptions of both groups have been unexplored. This study surveyed students with and without MHC in the same classes about their hypothetical disclosure of MHC. Students in several introductory biology classes were asked whether they identified as having MHC, whether they would or would not hypothetically disclose MHC to an instructor, and why. Thematic analysis identified reasons underlying their disclosure choices, which were sorted into the three beliefs of the Theory of Planned Behavior: attitudes, subjective norms, and behavioral control. Of the 803 respondents, 50% self-identified as having MHC. Students with MHC were less likely to say they would disclose their MHC to an instructor than students without MHC. Students with and without MHC who said ‘yes’ to disclosure gave similar reasons aligned with attitude beliefs. Students with MHC who said ‘no’ to disclosure perceived that the instructor wouldn’t care (attitude beliefs). Students without MHC who said ‘no’ to disclosure talked more about keeping their MHC private (subjective norms beliefs). Students without MHC who said ‘it depends’ talked more about impact on their course performance (attitude) than students with MHC. This research indicated that students with and without MHC do perceive disclosure differently and suggested that students with MHC focus more on negative instructor reactions, while those without MHC focus on privacy and performance. These differential perceptions may contribute to students with MHC seeing disclosure as a negative social cost versus a positive academic benefit.

## Introduction

A growing number of undergraduate students across the United States are being diagnosed with mental health concerns [1]. Mental health concerns (MHC) can encompass many different conditions with varying degrees of severity such as anxiety disorders, depression, post-traumatic stress disorder, and neurodevelopmental disorders, to name a few [2]. Anxiety and depression are some of the most common MHC experienced by undergraduate students. A national survey of multiple institutions found that 34.9% of undergraduates reported being diagnosed with anxiety and 27% reported being diagnosed with depression, including major depression, persistent depressive disorder, and disruptive mood disorder [3]. As student MHC increased during the pandemic, researchers found negative relationships with academic motivation and sense of belonging [4], suggesting negative impacts on student success.

Undergraduate students experiencing MHC are at an increased risk of negative outcomes in their courses and degree programs. Prior research at our institution found that biology students with higher levels of anxiety also reported a higher likelihood of leaving the biology major [5–6]. Research at other institutions found that students with depression and/or other MHC have less successful academic outcomes [7–8]. Given these negative impacts, students are often encouraged to disclose their MHC to their instructors to seek academic accommodations when they are needed, such as making up in-class work, extending assignment deadlines, or receiving make-up exams. However, these academic benefits can be seen by some students as having costs that weigh on students’ decisions to disclose.

MHC are one example of a concealed stigmatized identity (CSI), which has been associated with perceived risks to disclosure [9]. CSIs are identities that students may hold that can be hidden, but if revealed may change the way people view them and their abilities [9]. One study, for example, found that students with MHC did not want to request accommodations because they feared stigmatization by their teachers or other students [10]. A study by Busch and colleagues [11] documented that students with depression were concerned about being treated negatively or being viewed as making an excuse by their instructor. Thus, students with MHC must weigh the academic benefits of disclosure with the perceived personal costs when considering whether to disclose their needs and request an accommodation.

These disclosure costs and benefits are weighed by students in a classroom context that is increasingly evenly split between students with and without MHC [1,3]. And even though awareness about MHC is higher than in the past, almost half of students with MHC feel a perceived stigma from others [1]. Yet, studies on disclosure of MHC typically only focus on perceptions of disclosure by students with MHC and not the accompanying perceptions of students without MHC in the same contexts, making direct comparisons of perceptions difficult. To directly compare their perceptions, we asked students with and without MHC in several introductory biology classes about a hypothetical disclosure scenario to investigate whether beliefs about disclosure, including attitudes, subjective norms, and behavioral control, differed among these students. If these beliefs do differ, it confirms perceptions of differences by students with MHC and informs the types of barriers that students with MHC face as they weigh disclosure decisions.

### Factors impacting MHC disclosure

Institutions of higher education recognize that MHC can create barriers to student success. To lower these barriers, they encourage students to seek accommodations to reduce the impact of MHC on their academic outcomes. The process of getting an accommodation generally involves two steps: (1) working with the institutional disability office to document MHC needs, and (2) asking a professor for an accommodation [12]. As the number of students with MHC have risen in the college population, so has the number of students seeking institutional assistance [1,13]. Yet, there is still a large proportion of students who do not request or receive support for their MHC [13]. For some students, the difficulty of navigating the healthcare system to receive proper documentation of their MHC means they lack the ability to complete the official accommodation process [10]. Even with documentation, however, there are still many barriers to disclosure.

Some barriers that students face are logistical challenges related to institutional accommodation or treatment. Students in one study had challenges explaining what their needs were and therefore were unable to advocate for themselves to an institutional disability office; others did not think what would be offered would be helpful [14]. In a study that interviewed 33 engineering students, they articulated several challenges related to the institutional processes such as ease in accessing treatment, cost, timeliness, and getting the resources needed to treat their MHC [15]. Even after institutional approvals, the student still bears the responsibility of invoking the need for an accommodation with their professor [12]. These logistical challenges are not the only barrier to disclosure, however; many students also struggle with the way others might view their MHC.

Mental illness is one example of a CSI which include aspects of an individual’s identity that are not visibly identifiable and have negative stereotypes or attributes ascribed to them [16–18]. Despite increased awareness about MHC and some decline in perceived stigma, one study found 46% of surveyed students with MHC still perceived stigma about mental illness [1]. These perceptions of stigma may cause students to view disclosure as having a social cost because of fears of discrimination or disclosure reactions [16]. Another study found that students chose not to disclose their MHC to instructors or peers to distance themselves from negative stereotypes about MHC (such as others seeing them as not being capable), therefore maintaining a preferred social identity [19]. This indicates that despite the encouragement to disclose their need for accommodations and receive academic assistance, many students are weighing the benefits they might receive against the social costs disclosure might incur.

Two studies illuminate how college students with MHC weigh the potential support they might receive from disclosure with the concerns about negative impacts of that disclosure. Busch and colleagues studied how undergraduate students with depression viewed disclosure to their online biology instructors [11]. These students recognized that disclosure of MHC could increase student-instructor communication and add flexibility to their workload and deadlines. However, the perceived cost of disclosing MHC, such as being treated negatively or being viewed as making an excuse, outweighed the benefits for many students. Kranke and colleagues found that students often wanted to maintain autonomy and normality, which contributed to their reluctance to disclose MHC to a professor [20]. However, students were willing to talk about their MHC with a professor when they perceived the instructor was supportive and when they felt their academic performance could be jeopardized [20]. The work of these two studies provided empirical evidence that there are many factors that impact intentions to disclose MHC, and many of those perceived costs and benefits seem to be aligned with theoretical aspects of the Theory of Planned Behavior.

### Theoretical model

The Theory of Planned Behavior (TPB, Table 1) theorizes that the intent to enact a particular behavior is guided by individual beliefs about that behavior [21–23]. These beliefs include an individual’s attitudes, subjective norms, and behavioral control related to the behavior [22]. TPB is an ideal theoretical framework for this study as it is often used by researchers to understand how people make decisions about health-related behaviors [23]. In the context of our study, we are studying students’ intentions to disclose MHC to an instructor, which should be informed by students’ attitudes about disclosing MHC, the subjective norms they perceive about MHC, and their behavioral control beliefs about disclosing MHC (Table 1). Importantly, this theory can be applied to students with and without MHC in our study because we are presenting them with a hypothetical scenario and asking them about their intent to disclose in that situation.

**Table 1.** The Three Beliefs of Ajzen’s Theory of Planned Behavior. The three beliefs guiding the intention to act are shown with examples specific to disclosing MHC to an instructor. These beliefs inform the intention to disclose to an instructor, which is aligned theoretically with the behavior of disclosing.

| TPB belief | Beliefs, as contextualized in this study |
| --- | --- |
| Attitudes | How does the student feel about disclosing MHC to an instructor? |
| Subjective Norms | How do others perceive the disclosure of MHC? |
| Behavioral Control | How comfortable is the student with disclosure of MHC? |

‘Attitude’ beliefs are student perceptions about how *they* might feel (positively or negatively) about disclosing MHC or their perception about the positive or negative outcomes that might arise from the disclosure. A student may believe, for example, that disclosing MHC may result in a flexible deadline for an assignment (a positive outcome), while another student may perceive that they would feel shame in disclosing their MHC (negative feeling). ‘Subjective norms’ are beliefs about how an individual thinks *others* will view their MHC disclosure or how others they respect might act in a similar situation. Thus, a student would have a higher intention to disclose MHC if those they respected in their social network (i.e., friends, family, peers) viewed disclosure as a positive and/or acceptable behavior. However, they might be less inclined to disclose if those in their social network do not disclose MHC despite the need to do so. Finally, ‘behavioral control’ is the extent to which an individual thinks they have the agency to disclose MHC, based on their confidence and perceived barriers. A student with high self-confidence about their ability to disclose MHC to an instructor would be more likely to do so. A student who perceives significant situational barriers to disclosure would be less inclined to do so.

A recent study used TPB to investigate MHC disclosure to instructors among 311 students in a communications course at a small private California University [24]. They found that all three TPB beliefs were related to intention to disclose, but subjective norms beliefs were most strongly related to that outcome. They did not disambiguate the results by those with and without MHC, however. Their findings support that attitude, subjective norms, and behavioral control beliefs are perceptions a student with or without MHC would use to react to the scenario about intention to disclose MHC in this study. Even though beliefs vary by individual, there may be patterns in how particular populations (e.g., students with and without MHC in introductory biology classes) relate beliefs to particular intentions (e.g., disclosing MHC). Therefore, we investigated which of the beliefs might drive intent to disclose MHC in our introductory biology population, and whether those beliefs differed among students with and without MHC, to explore potential differences in student perceptions about MHC and its disclosure that might indicate the potential for stigma.

### Rationale

Prior research provided a foundation for investigating the costs and benefits students with MHC perceive when they are weighing disclosure of MHC to an instructor [11,20]. However, one study was specific only to depression and disclosure in an online setting [11], and both studies only probed the perceptions of students with MHC. Our study asked students with and without MHC about their hypothetical intentions to disclose MHC to an instructor and used the reasons for their disclosure choices to determine their TPB beliefs. We posited that students with and without MHC would make different choices about intention to disclose that were driven by different TPB beliefs, with students with MHC mentioning subjective norms more often due to the stigmatized nature of MHC. To explore this hypothesis, we collected data to answer the following research questions:

1) Do students with and without MHC make different choices about hypothetical disclosure of MHC to an instructor?
2) What are the differences in disclosure reasoning for students with and without MHC?

If disclosure intentions and TPB subjective norms differ between students with and without MHC in introductory biology classes, it would potentially support perceptions of stigma that impact whether students with MHC disclose support needs to their instructors. Our findings suggested evidence for the existence of a stigmatized environment related to MHC disclosure and highlight a need for instructors to create a classroom climate that decreases these perceptions in order to encourage support for all students.

## Methods

### Courses and participants

This study surveyed students in two large introductory biology courses at one research-intensive university in the southeastern United States. One of the courses was introductory biology for non-science majors focused on molecules, cells, and body systems, and the other was introductory biology for majors focused on ecology, evolution, and biodiversity. The non-science majors’ course is taken as a general education credit by students in many majors, but is also an option for students in nursing, kinesiology, and psychology. The majors’ course is the first in a two-course sequence and is typically composed of about 70% STEM majors and 30-40% biology majors.

Surveys were sent to students in five lecture sections of the non-science majors’ course and two lecture sections of the science major’s course. All the instructors were non-tenure track faculty with several year’s experience teaching introductory biology courses. There were three non-science major lecture sections with 225 students and two with 150 students for a total of 975 students in that course. One of the science major sections had 200 students and one had 225 students for a total of 425. This study was approved under institutional review board protocol #23-07777-XP.

### Data collection

The data for this study were collected during the fall 2023 semester. The researchers provided instructors of the classes a recruitment email with survey link and consent information and asked them to forward the information to their students. Students clicked the link to the survey, read the consent information, and then clicked yes or no to provide permission to use their data for the research study (as consistent with a waiver of written consent from our institutional review board). The survey was distributed to students on November 20, 2023 and was available until December 5th, 2023. Instructors were allowed to incentivize participation by offering one point of class credit; classes had 800-1,000 points total. We utilized Qualtrics Survey software (https://www.qualtrics.com) as the surveying platform.

### Survey instrument

The survey questions (S1 Table) were developed by the authors and reviewed by education research experts for clarity and alignment with the research questions. Students were first asked, “Do you currently identify as having a mental health concern (whether documented or not), e.g., anxiety, generalized anxiety disorder, depression, bipolar disorder, PTSD, panic disorder, suicidal ideation?” to which they answered yes, no, or prefer not to answer. Students were then asked, “Hypothetically or not, if you were experiencing a mental health concern that was or could be impacting a course task (e.g., meeting a course deadline and/or studying for an exam and/or taking an exam), would you disclose the mental health concern to your instructor? Explain Why.” They could answer yes, no, or it would depend. In addition to those questions, there were questions about course type (non-science major or science major course), race and/or ethnicity, college generation status, year in college, gender, and major (answer choices for all demographic questions provided in the S1 Table) that were used solely to describe the sample.

### Data analyses

Partial responses were removed from the dataset, as well as data from students who were not 18, who did not give consent for their responses to be used in the study, and who indicated that they preferred not to respond to the disclosure question. After removing these data, the final sample size was 803 students.

#### Do students with and without MHC make different choices about hypothetical disclosure of MHC to an instructor?

We sorted students into those who did or did not identify as having MHC and summed the disclosure choice (yes, no, or prefer not to answer) for each group. Percentages were calculated for each, and a chi-square test was conducted to identify whether there were significant differences in the distribution of disclosure choices between students with and without MHC [25]. Significance was indicated by a p-value less than alpha which was equal to 0.05.

#### What are the differences in disclosure reasoning for students with and without MHC?

Students followed up their disclosure choice with a written explanation describing the reason for their choice. Two coders (M.S. and E.S.) inductively analyzed student responses to create categories of responses that could be named and described, in a process called thematic analysis [26], and then these codes were matched to the TPB beliefs in a deductive process. E.S. has conducted biology education research on introductory biology classes for over 15 years, including work on the impacts of anxiety. M.S. is an undergraduate researcher interested in mental health and well-being who worked with the research lab of E.S. to undertake this work. B.V.D.M. is a graduate student in the lab of E.S. who also conducts research on introductory biology classes; she reviewed the process of deductively sorting the codes. The researchers met weekly through the design, data collection, and analysis of the study. They closely monitored and discussed their interpretation of the data during the analysis phase.

Inductive coding was chosen because the research team wanted the codes to reflect student voice, versus forcing student voice conform to theory (i.e., using TPB to deductively code our data). Only after codes were inductively identified were they aligned with TPB beliefs. To conduct thematic analysis, the two coders started by independently reading the responses and taking notes on common ideas mentioned by the students. The researchers then discussed their notes to create an initial codebook, which is a tool to guide the assignment of student responses to specific categories. Codebooks are iteratively refined through rounds of testing by the coders until they have evidence, they have reached a minimum agreement percentage on the assignment of codes; for us, this minimum was set at 70% [27]. In this study, the researchers went through three rounds of independently coding 50 student responses, discussing the responses, and revising the codebook before they felt comfortable moving into the final coding phase.

For final code assignment, half of the student responses (n=400) were coded independently by M.S. and E.S. Using the codebook, one student response was considered the coding frame and was assigned as many codes as needed to represent the ideas being conveyed by the student. Agreement among codes (as assessed by individual codes and not the entire unit) assigned to the responses was 72% for this set of responses. The coders then met to discuss the codes they did not agree on and revised coding decisions to reach 100% agreement. As they worked, they made minor edits to the codebook descriptions and added more examples. Having met the standard of 70% agreement, E.S. then worked independently to code the remaining 403 student responses using the newly refined codebook that consisted of ten codes.

To align the codes identified through the inductive analysis with the three beliefs of TPB (attitude, subjective norms, behavioral control), all authors read the theoretical descriptions of each belief and independently placed each code from the codebook into a belief group. After viewing and discussing this initial sorting, the research group decided to create belief prompts to aid in the placement of the codes. For attitude beliefs, the prompts were: (1) Disclosure could make me feel and (2) If I disclose, this could happen. For subjective norms beliefs the prompts were: (1) Other people think that disclosing is and (2) People I know. For behavioral control beliefs, the prompts were: (1) I feel about my capability to disclose or (2) Disclosure will be easier or harder because of. The group used these prompts to sort the codes again and discussed their placement to consensus. The team agreed that there were two inductive codes that could be aligned with two TPB beliefs; these were called “hybrid” codes.

To answer the second research question, the data were sorted into students with and without MHC, and then each of these groups was subdivided based on their disclosure intention: yes, no, or it would depend. Within each of these six groups, the percentage of students who mentioned each of the codes was calculated by summing the code incidence and dividing by the number of students in the group. In presenting these data, we highlighted >10% differences in code prevalence between students with and without MHC and removed any codes with less than 5% prevalence across all six groups.

## Results

There were 803 respondents to the survey (Table 2). Of these, 422 were from the non-science majors’ course (43% response rate) and 381 were from the science majors’ course (90% response rate). The sample overall was majority women (76.6%), white (81.7%), continuing generation (85.6%), first-year students (66.3%). Across the entire sample, 50% of the respondents self-reported MHC; with 49% of the students in the non-science majors course indicating MHC and 50% in the science majors’ course.

**Table 2.**
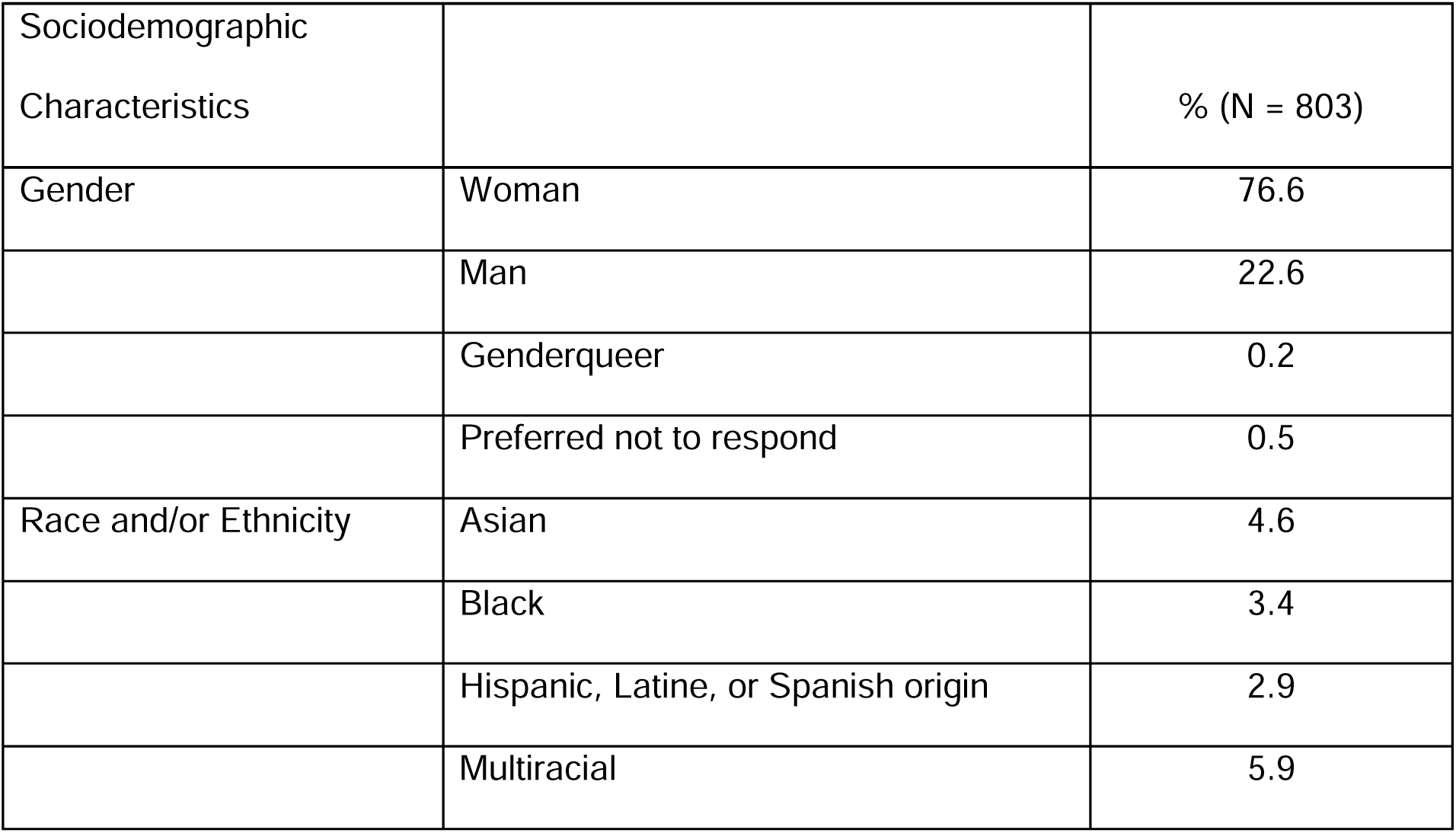
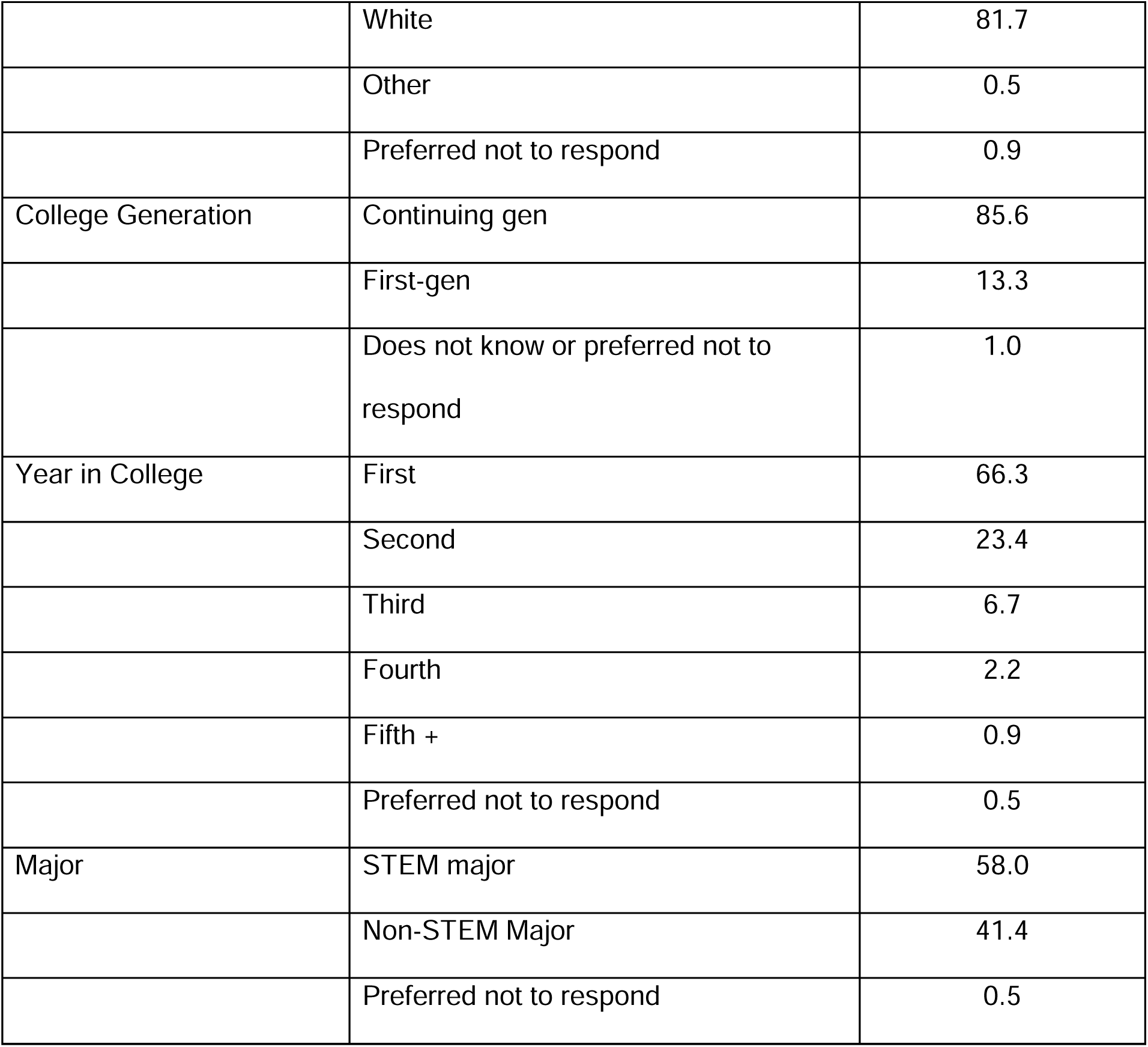
Demographic characteristics of the survey respondents.

| Sociodemographic Characteristics |  | % (N = 803) |
| --- | --- | --- |
| Gender | Woman | 76.6 |
|  | Man | 22.6 |
|  | Genderqueer | 0.2 |
|  | Preferred not to respond | 0.5 |
| Race and/or Ethnicity | Asian | 4.6 |
|  | Black | 3.4 |
|  | Hispanic, Latine, or Spanish origin | 2.9 |
|  | Multiracial | 5.9 |
|  | White | 81.7 |
|  | Other | 0.5 |
|  | Preferred not to respond | 0.9 |
| College Generation | Continuing gen | 85.6 |
|  | First-gen | 13.3 |
|  | Does not know or preferred not to respond | 1.0 |
| Year in College | First | 66.3 |
|  | Second | 23.4 |
|  | Third | 6.7 |
|  | Fourth | 2.2 |
|  | Fifth + | 0.9 |
|  | Preferred not to respond | 0.5 |
| Major | STEM major | 58.0 |
|  | Non-STEM Major | 41.4 |
|  | Preferred not to respond | 0.5 |

### Do students with and without MHC make different choices about hypothetical disclosure of MHC to an instructor?

For the hypothetical question of whether MHC would be disclosed to an instructor, student responses (yes, no, or it would depend) differed significantly between students with and without self-reported MHC (□²(2, *N* = 803) = 53.47; p < 0.001). Students with and without MHC were almost identical in their selection of the response ‘it would depend’ (56% and 57%, respectively) (Table 3). However, those who self-identified as having MHC were less likely to say they would disclose MHC (‘yes’) to an instructor (9%) compared with those without MHC (25%). For those who self-identified as having MHC, 35% said they would not disclose to their instructor, while 18% of those without MHC said they would not disclose to an instructor.

**Table 3.** Comparison of disclosure choices between students with and without self-reported MHC.

|  | Yes MHC (50%, *n*=398) | No MHC (50%, *n*=405) |
| --- | --- | --- |
| It would depend | 56% (*n*=223) | 57% (*n*=231) |
| Yes | 9% (*n*=34) | 25% (*n*=100) |
| No | 35% (*n*=141) | 18% (*n*=74) |

### What are the differences in disclosure reasoning for students with and without MHC?

#### Disclosure codes

The inductive codes from student responses were sorted into the three TPB belief categories and one hybrid category: 1) Attitude beliefs, 2) Subjective Norms, 3) Behavioral Control beliefs, and 4) Hybrid beliefs (Table 4). There were five inductive codes in the attitude beliefs category, two in the subjective norms category, one in the behavioral control beliefs category, and two in the hybrid beliefs category. Brief descriptions of each TPB belief and code, along with an exemplar student quote, are shown in Table 4.

**Table 4.** Inductive codes generated from student responses, organized into TPB beliefs. The codes represent common categories about why students would or would not disclose MHC to an instructor.

| TPB belief category and inductive codes | Exemplar student quote for each code |
| --- | --- |
| ATTITUDE: how a student felt about disclosing or about the disclosure outcome.<br>Prompts: (1) Disclosure could make me feel ____ and (2) If I disclose, this could happen ____.<br>1. Embarrassing - student said disclosing would make them feel ashamed or nervous; said it might make MHC worse because of increased anxiety.<br>2. Instructor won't help - student said instructor likely wouldn't provide help or accommodation, so there was no use in disclosing; some said it may be because they lacked documentation or diagnosis to validate MHC. | *"I know that many instructors are very understanding, but letting one know you need help or are behind because of mental health almost feels embarrassing."*<br>*"I feel like a lot of professors would not really do anything about it."* |
| 3. Instructor aware and might help -<br>Student said they would disclose to instructor early in the semester in case they needed help later; thought instructor would help, support, accommodate; they would disclose because mental health is important.<br>4. Impact on performance - student said they would disclose if it was impacting success, learning, or attendance in a class.<br>5. Instructor may or may not care - student said it would depend on whether they thought the instructor would believe them / think the concern was valid; or whether they thought the instructor would care, understand, show concern and respect, listen, and take them seriously. | *"I would inform each of my professors about it so we could hopefully do what it takes to get around it."*<br>*"It would depend on how much it was impacting the class."*<br>*"But sometimes instructors don't take that seriously and say that you should've known you would struggle and should've come to them sooner..."* |
| SUBJECTIVE NORMS: how a student thought others might view their disclosure or |  |
| act themselves. Prompts: (1) Other people think that disclosing is ____ and (2) People I know ____.<br>1. Private to student - student would prefer not to disclose because it is a personal concern, not the instructor's; student said they would not be comfortable talking about it.<br>2. Perceptions others have - student wouldn't want to disclose because others may view an accommodation as an excuse or special treatment and then see you differently or "less than" in some way. | *"I typically think of this as private information that is not to be discussed."*<br>*"I would tell my teacher, but would try to meet in person so that they didn't think it was an excuse and that I was slacking off on that assignment."* |
| BEHAVIORAL CONTROL: how a student felt about their abilities or barriers to disclosure. Prompts: (1) I feel ____ about my capability to disclose or (2) Disclosure will be easier or harder because of ____.<br>1. Approachability of instructor - student | *"Some professors are more open to helping* |
| said it would depend on how comfortable they were with instructor, how open the instructor is to talking with students. | *students...”* |
| HYBRID: combination of two of the TPB belief categories<br>1. Severity of MHC - student said they would only disclose if it was having a major impact on attendance; if it was an illness with broad impacts; they often talked about this as a last resort.<br>(hybrid of attitude / control)<br>2. Won't let it impact - student said there would be no need to disclose because they wouldn't let it impact their performance or they would deal with the consequences on their own.<br>(hybrid of subjective norms or control) | *“It would depend on what I am going through, as some situations can be more severe than others.”*<br>*“I have had issues before and personally I feel that getting used to them, doing what you can to help, or simply dealing with the consequences is something that has to be done on your own.”* |

ATTITUDE codes were beliefs students had about how they might feel if they disclosed their MHC to an instructor (embarrassing) or what outcome they might expect (instructor won’t help, instructor aware and might help, impact on performance, instructor won’t care). The code ‘embarrassing’ was the perception that having to disclose MHC might make the student feel ashamed or nervous. A few students mentioned that disclosing could make their MHC worse because of the anxiety it would cause them. The code of ‘instructor won’t help’ was when students said they would not disclose to an instructor because they did not think the instructor would do anything; students mentioned that this might be because of strict course policies or their lack of MHC documentation. The code of ‘instructor aware and might help’ was when students said they should disclose their MHC to their instructor because MHC advocacy was important, and the instructor needed to know about their concerns. Many students said specifically that this was important because the instructor would then work to provide resources or assistance to help. The ‘impact on performance’ code was a risk / benefit assessment the student was making to decide how much class work they could miss before they would need to ask an instructor for help. The code of ‘instructor won’t care’ was based on student perceptions of negative reactions by the instructor in response to their disclosure. Some students talked about the instructor not believing that the MHC was real or that it would cause a problem; some said that the instructor would not show care or respect or take their concern seriously once they disclosed the MHC.

SUBJECTIVE NORMS codes were beliefs students had about how others might view their disclosure or what others they respected might do in the same situation. The code of ‘private to student’ was when students talked about MHC being a personal concern that they would not want to disclose to an instructor. Students often talked about the comfort level of disclosure related to privacy. The code of ‘perceptions others have’ was when students talked about whether disclosure and subsequent accommodation might result in them being viewed by their peers or instructor differently because of the special treatment they had received; they were particularly concerned about it making them appear “less than” others.

BEHAVIORAL CONTROL were beliefs about conditions that made it easier or harder for the student to disclose, or their capability to disclose. The only code in this category was ‘approachability of instructor’ which was when students talked about how open the instructor seemed to be with regards to talking with students about concerns like MHC.

There were two HYBRID codes which were student beliefs that seemed to cut across two of the belief categories above. The code of ‘severity of MHC’ was when the participant talked about their decision to disclose depending on the type of MHC or how severe the MHC was because that would determine how much school they might miss. This could have been based on a perceived outcome, but students also talked about this being out of their control, so they lacked agency. The code of ‘won’t let it impact’ was when students said they would not need to disclose MHC because it was not something to be used as an excuse to not do their work; they would do the work regardless of the MHC. On one hand this was showing agency, but there was also a hint of not wanting to disclose which could reflect perception of a norm.

#### Patterns in disclosure explanations

The prevalence of each code, aligned with their TPB belief categories and sorted by disclosure choice and MHC status, is shown on a heat map (Fig 1). This allows for two interpretations: (1) which codes and TPB beliefs are driving disclosure choices overall and (2) how students with and without MHC reason may differ in their disclosure reasoning.

**Fig 1.** Heat map of disclosure reasoning prevalence by student groups. The heat map shows the relative prevalence of codes explaining student reasoning for their disclosure choices, sorted by disclosure choice and whether they do or do not have MHC. The codes are aligned by the TPB belief categories on the left. Percent prevalence of codes are increasingly darker with higher relevance, as shown by the key on the right.

*Disclosure explanations overall –* For each disclosure choice (yes, no, it would depend) there was one most prevalent code. Students who said they would disclose to an instructor mostly talked about the need to proactively make the instructor aware of their needs, which was a TPB attitude belief. For example, one student said, “As someone with anxiety, I always disclose my health concerns with my professors, as I get extra time for tests and quizzes. I let them know at the beginning of the semester” Students who said they would *not* disclose to an instructor mostly talked about the subjective norms belief of privacy. One student said, “I feel like society does not view it as a valid reason to miss an assignment.” Students who selected ‘it would depend’ as their disclosure choice mostly talked about performance impacts, an attitude belief. One student, for example, said, “I would always disclose this with my professor if I felt it was impacting my performance. My academic success is my priority and it is important to be open if something could be interfering with that.” Although these codes were the most commonly voiced for each disclosure choice, there were differences between students with and without MHC.

*Disclosure reasoning between students with and without MHC –* For students who said they **would disclose** to an instructor, students with and without MHC had similar code percentages suggesting similar beliefs about their disclosure choice. For students who said they **would not disclose**, we highlight three differences. Students without MHC were more likely to say that MHC were private to them (47.30%) in comparison to students with MHC (34.75%). Conversely, students with MHC were more likely to say that the instructor would not care or believe them (21.28%) compared to only 10.81% of students without MHC. One student with MHC voiced, “Many professors do not understand mental illnesses or simply are not concerned about it.” Finally, students with MHC were also more likely to say that the instructor would not help them (17.02%) compared to students without MHC (6.76%). As one student with MHC indicated, “I have always seen these requests ignored despite having a professional diagnosis.” For students who said **disclosure “would depend,”** there was only one code difference: students without MHC were more likely (35.93%) than students with MHC (25.11%) to mention that an impact on their performance was important to their disclosure.

## Discussion

Collegiate courses are increasingly an equal mix of students with and without MHC [1,4]. In the introduction we posited that students with and without MHC in introductory biology courses would view disclosure intention differently and have different beliefs about subjective norms that may indicate a stigma associated with disclosure for students with MHC. Our prediction that students with and without MHC would have different disclosure intentions was affirmed; students with MHC were less likely to say they would disclose an MHC to an instructor. We studied the reasons behind these disclosure choices and found that students who said they would disclose had similar reasons regardless of MHC status. The reasons why students would not disclose or were uncertain about it differed across MHC status in ways we did not predict. Those with MHC expressed more concerns about how an instructor would negatively react to their disclosure (attitude belief), while those without MHC were more concerned about privacy (subjective norms belief) or performance impact (attitude belief). These findings provided evidence that students with and without MHC in the same classes perceive disclosure differently and suggest that attitude and subjective norms beliefs drive these differences [21–22]. Students without MHC being concerned about keeping MHC private and students with MHC being concerned about instructor reactions suggests the reality that stigmas exist in these introductory biology classes and may temper the climate for students with MHC.

### Do students with and without MHC make different choices about hypothetical disclosure of MHC to an instructor?

In our study, students with MHC were less likely to say they would disclose to an instructor compared to their peers without MHC. Others have documented a reluctance by students with MHC to disclose, even in the face of increasing support and awareness about MHC [13]. However, our study is the first, to our knowledge, to directly compare differences in disclosure intentions between students with and without MHC. Interestingly, we found that more than half of students with *and* without MHC indicated that their disclosure “would depend” on other factors. This indicates that students without MHC can envision barriers and benefits to disclosure of MHC, although our results suggest they may be doing so differently from students with MHC. Almost 10% of students with MHC said they would disclose. Some research has suggested that prior disclosure makes it more likely for a student to say they would disclose [24], however, the nature of these prior disclosure experiences matters, with negative prior experiences leading to increased reluctance to disclose in the future [28]. Given this, studies focused on how prior disclosure impacts future disclosure would help understand the extent to which those experiences are contributing to perceptions of stigma in introductory classrooms.

### What are the differences in disclosure reasoning for students with and without MHC?

#### Attitude and subjective norms beliefs were most prevalent in shaping disclosure perceptions

We aligned our codes of students’ rationale for disclosure with the Theory of Planned Behavior (TPB) [21–23]. We found that for our prompt and this population, attitude and subjective norms beliefs were most salient to students’ intention to disclose. Attitude beliefs were most often related to the intention to disclose and to the potential positive outcomes from working with a professor to accommodate MHC disruptions or revealing an MHC when course performance was being impacted. These results suggest the most prevalent benefit students were weighing was the motivation to perform well in these courses. Subjective norms beliefs were most commonly related to a perception that MHC were private and were one of the leading rationales cited by students who said they would not disclose. Given that the disclosure prompt was a scenario about MHC impacting course performance, about a third of students were willing to suffer academic consequences for the sake of privacy. Our results differed slightly from the recent quantitative analysis relating TPB to instructor disclosure intentions; they found that all three beliefs were related to intention to disclose, with subjective norms being most strongly related [24]. However, their data collection involved forced choice responses versus our open-ended prompts, making the results difficult to compare. The disclosure climate at a small private college in California may also be significantly different than at a large research-intensive institution in the southeast. We suggest that there is a need for more research on MHC disclosure guided by TPB. Specific to our study, we see a need to explore cost / benefit tradeoffs between attitude and subjective norms beliefs such as performance and privacy.

These could be explored using scenarios with various academic impacts and accommodations to better understand when students are willing to break norms for potential positive outcomes.

#### Disclosure reasoning differed by disclosure choice and MHC status

We found few differences in disclosure reasoning among students who said yes to disclosure, regardless of MHC status. These students often indicated proactive help-seeking, talking about their intention to disclose their MHC to the instructor in advance of any problems so they could work together in the future. Very few of these students talked about concerns related to privacy. This mirrors findings by Busch and colleagues [11] that some students saw the benefits of building relationships with professors to support MHC. Although disclosure reasoning was similar among those with and without MHC, there were far fewer students with MHC who said yes to disclosure (9% of students with MHC compared to 25% without MHC), and disclosure overall was low across both groups. This aligns with reports of low rates of disclosure even as MHC support has increased [13], but also indicates that even students without MHC are reluctant to disclose. Interviewing students who are willing to disclose may provide more insight into whether they see fewer barriers, whether they see them differently, or how they cope with disclosure concerns that may inform work on interventions to increase disclosure.

Students with and without MHC differed in disclosure reasoning when it came to why they said ‘no’ or ‘it would depend.’ For students with MHC, the perception that instructors may not care and the belief that the instructor would not help following disclosure was a significant hindrance to disclosure. Similarly, Busch and colleagues [11] also found that students with depression feared negative reactions from their instructors. Research has found that students who had had negative disclosure experiences in the past were less likely to view future disclosure positively [28], suggesting that past experiences could have driven perceptions in this study as well. Previous studies also found that students weigh stigma and fear of negative consequences when deciding whether to disclose MHC to instructors [11,17–18,20], and while our study found this as well, these perceptions were higher in students without MHC than those with MHC. As the incidence of MHC have risen in the student population over time, awareness has also risen about them, and there has been some reduction in perceived stigma among those with MHC [1]. Our study may have reflected these changes, with students with MHC voicing fewer subjective norms concerns compared with those without MHC; this may be because institutional accommodation processes already required them to reveal MHC, making it more of a norm for them [12]. However, the findings that those without MHC were reluctant to disclose because of subjective norms beliefs hints at the continuing negative perceptions about MHC held by peers of students with MHC. Overall, the reasons why students might be hesitant to disclose MHC hint at the ways in which students with MHC navigate in a class environment where they do not always trust instructors to help and have peers who may not understand their concerns.

### Supporting students in introductory biology with MHC

Half of the students in each class we surveyed self-reported MHC. These results reflect the continuing rate of mental health challenges among undergraduate students [1,3] and are of concern because of the documented negative impacts MHC can have on student success [5–8]. Although there are many institutional units to support these students, instructors play an important role in how they create classroom climates of support, inclusion, and belonging.

Instructors can build a positive climate early through inclusive policies and syllabi that communicate empathy and support [29–30]. Classroom communication that welcomes feedback, promotes dialogue, and conveys approachability and relatability may help students feel comfortable disclosing concerns because it conveys that the instructor is caring [31]. Anti-stigma interventions [32] might also be considered as ways to signal openness to hearing student concerns, but also build understanding across students with and without MHC in a class. It is also likely that there are different rates of student disclosure among different introductory biology courses; future research could identify courses where students feel more comfortable disclosing and identify features that could be emulated in other courses. Care needs to be taken, however, to support instructors who are already experiencing their own levels of burnout and MHC so as not to add additional workload that could decrease their effectiveness.

### Limitations and future work

The sample for this study was drawn from a single institution, constraining our ability to generalize to the larger population of students at other institutions. This study was also conducted only in introductory biology courses, which are a unique sub-population at our institution and further bound any inferences we can make. Studies in other classes and at other institutions, particularly with a more representative sample of students from historically marginalized groups, would provide additional evidence about whether the differential student reasoning for disclosure are generalizable to a broader population.

This study focused on students’ intent to disclose versus actual disclosure. Although theory suggests a tight relationship between the two, actual disclosure was not studied, nor could it be given the lack of MHC in half the respondent population. Asking students without MHC to imagine a scenario where they have MHC may not represent how they would actually act. However, our results do hint at the differential assumptions held by each group, which highlight a need to better understand not just reasoning behind disclosure, but also where those ideas originate. A study on this could probe the relative impacts of prior disclosure, perceptions of stigma, motivation to perform well in a course, and instructor trust [33], among others.

## Conclusion

Given the percentage of students with MHC at undergraduate institutions [1] and the negative impacts MHC can have on their collegiate performance, it is important to understand how their unique perspectives and experiences may differentially impact their success. This study suggests that students with MHC have different perceptions compared to their peers without MHC about disclosing an MHC to an instructor. Students with MHC indicated negative perceptions of instructor reactions to disclosure that could act as barriers to help-seeking. Their peers without MHC were concerned more about privacy and performance when considering the same hypothetical disclosure question. This suggests students with MHC navigate in an environment where they may assume less understanding from their instructors and interact with peers who think MHC should be private. Future research should focus on factors to bridge these classroom perceptual differences to ensure an environment where students with MHC may disclose their needs without fear of negative judgment or inadequate assistance.

## Data Availability

Data produced in the present work are contained in the manuscript.

## Acknowledgements

We thank the instructors and students who helped us collect data, feedback on the manuscript from Dr. Maryrose Weatherton and Hope Ferguson, and inspiration for the project from Drs. Kelly McDonald and Cathy Ishikawa.

## Supporting information

**S1 Table.** Survey questions and associated purpose for the study.

|  | Would Disclose? | Yes |  | No |  | Maybe |  |
| --- | --- | --- | --- | --- | --- | --- | --- |
|  | MHC? | Yes | No | Yes | No | Yes | No |
| TPB Theme | Code |  |  |  |  |  |  |
| Attitude |  |  |  |  |  |  |  |
|  | Embarrassing | 0 | 1.00 | 5.67 | 10.81 | 7.62 | 3.03 |
|  | Instructor won't help | 2.94 | 0 | 17.02 | 6.76 | 9.42 | 3.46 |
|  | Instructor aware and might help | 64.71 | 71.00 | 0 | 1.35 | 4.93 | 9.09 |
|  | Impact on performance | 38.24 | 39.00 | 3.55 | 0 | 25.11 | 35.93 |
|  | Instructor may/may not care | 5.88 | 7.00 | 21.28 | 10.81 | 17.49 | 7.79 |
| Subjective Norms |  |  |  |  |  |  |  |
|  | Private to student | 5.88 | 0 | 34.75 | 47.30 | 18.83 | 16.02 |
|  | Perceptions others have | 2.94 | 0 | 17.02 | 12.16 | 8.97 | 4.33 |
| Behavioral Control |  |  |  |  |  |  |  |
|  | Approachability of Instructor | 2.94 | 9.00 | 7.09 | 5.41 | 10.31 | 14.72 |
| Hybrid |  |  |  |  |  |  |  |
|  | Severity of MHC | 0 | 1.00 | 0 | 2.70 | 19.28 | 23.81 |
|  | Won't let it impact | 0 | 0 | 19.86 | 13.51 | 13.90 | 7.36 |

## Notes

### Competing Interest Statement

The authors have declared no competing interest.

### Funding Statement

This study did not receive any funding

### Author Declarations

IRB of The University of Tennessee gave ethical approval of this work.IRB-23-07777-XP

