## Supplemental Table 1 for "Undergraduate students with and without mental health concerns have different perceptions of disclosing mental health challenges to instructors"

S1 Table. Survey questions and associated purpose for the study.

| **Survey question** | **Purpose** |
| --- | --- |
| **STATEMENT OF CONSENT** I have read this form and the research study has been explained to me.  I have been given the chance to ask questions and my questions have been answered.  If I have more questions, I have been told who to contact. By selecting “agree” below, I am agreeing to be in this study. If I select “do not agree” I will receive the credit for the survey, but my responses will not be used by the research team.  I agree  I do not agree | Consent statement |
| Please enter your age  (open-ended) | Screening for respondents over 18 and able to provide consent |
| Please select the course associated with this survey.  Bio 101  Bio 150  Bio 160 | Organizing participants into non-majors (101) or majors courses (150 or 160) |
| Do you currently identify as having a mental health concern (whether documented or not), e.g., anxiety, generalized anxiety disorder, depression, bipolar disorder, PTSD, panic disorder, suicidal ideation?  Yes  No  Decline to answer | Organizing responses into those with or without self-disclosed MHC |
| Hypothetically or not, if you were experiencing a mental health concern that was or could be impacting a course task (e.g. meeting a course deadline and/or studying for an exam and/or taking an exam), would you disclose the mental health concern to your instructor?  Yes, I would always disclose this concern  It would depend on the circumstances  No, I would never disclose this concern | Organizing responses into disclosure intention |
| Why did you answer the way you did above?  Open response | Generating disclosure reasoning responses |
| With which racial/ethnic group(s) do you identify? Choose all that apply  American Indian or Alaska Native  Asian  Black or African American  Middle Eastern or North African  Hispanic, Latino, or Spanish origin  Native Hawaiian or other Pacific Islander  White  Another race or ethnicity not listed above (please specify)  I do not wish to disclose this information | Demographic information to describe sample |
| Are you the first in your family to go to college?  Yes  I don’t know / prefer not to answer  No | Demographic information to describe sample |
| What year are you in college?  First-year  Second-year  Third-year  Fourth-year  5^th^-year or higher | Demographic information to describe sample |
| How would you describe your gender identity?  Woman  Man  Genderqueer  Agender | Demographic information to describe sample |
| What is your major?  STEM major (please specify)  Non-STEM major (please specify) | Demographic information to describe sample |
